# Safely return to schools and offices: early and frequent screening with high sensitivity antigen tests effectively identifies COVID-19 patients

**DOI:** 10.1101/2021.10.08.21264782

**Authors:** Yong Dam Jeong, Keisuke Ejima, Kwang Su Kim, Shoya Iwanami, Shingo Iwami, Kazuyuki Aihara

**Author notes:** Corresponding authors (KE) (S Iwami). These authors contributed equally to this work (Yong Dam Jeong, Keisuke Ejima, Kwang Su Kim). **Competing Interest Statement:** The authors declare that they have no competing interests.

## Abstract

**Background:** In-person interaction at school and offices offers invaluable experience to students and benefits to companies. In the midst of the pandemic, ways to safely go back to schools and offices have been argued. Centers for Disease Control and Prevention (CDC) recommends taking all precautions such as vaccination and universal indoor masking. However, even if all the precautions are implemented and transmission is perfectly prevented in the facilities, they may be infected outside of the facilities, which would be a source of transmission in the facilities. Therefore, identifying those infected outside of the facility through screening is essential to safely go back to schools or offices. However, studies investigating the effectiveness of screening are limited. Further, it is not well clarified now which screening strategy (e.g., low or high sensitivity antigen tests, intervals and the number of tests) effectively identify infected and infectious individuals to avoid transmission in facilities

**Methods:** We assessed the effectiveness of various screening strategies in schools and offices through quantitative simulation. The effectiveness was assessed by the proportion of identified infected and infectious participants. Infection dynamics in the facility is governed by transmission dynamics of the population they belong to, and the screening is initiated at different epidemic phases: growth, peak, and declining phases. The viral load trajectory over time for each infected individual was modelled by the viral dynamics model, and the transmission process at the population level was modelled by a deterministic compartment model. The model parameters were estimated from clinical and epidemiological data. Screening strategies were varied by antigen tests with different sensitivity and schedules of screening over 10 days.

**Results:** Under the daily screening, we found high sensitivity antigen tests (the detection limit: 6.3 × 10^4^ copies/mL) yielded 88% (95%CI 86-89) of effectiveness by the end of 10 days screening period, which is about 20% higher than that with low sensitivity antigen tests (2.0 × 10^6^ copies/mL). Comparing screening scenarios with different schedules, we found early and frequent screening is the key to maximize the effectiveness. Sensitivity analysis revealed that less frequent tests might be an option when the number of antigen tests is limited especially when the screening is performed at the growth phase.

**Discussion:** High sensitivity antigen tests, high frequency screening, and immediate initiation of screening are the key to safely restart educational and economic activities allowing in-person interactions. Our computational framework is useful in assessment of screening strategies by incorporating additional factors for specific situations.

## Introduction

SARS-CoV-2, the causative virus of COVID-19, quicky spread over the world in early 2020. One of main reasons why most of the countries failed containment of COVID-19 are its high transmissibility (*R*_0_ *=* 2.9) (Billah, Miah, & Khan, 2020) and substantial pre-symptomatic and asymptomatic infections (Arons et al., 2020; Kimball et al., 2020; Zhen-Dong et al., 2020). Although massive vaccination campaign is ongoing, substantial population are not vaccinated and/or not willing to be vaccinated (Malik, McFadden, Elharake, & Omer, 2020; Solís Arce et al., 2021). Thus it may be difficult that the vaccination coverage reaches the level of herd immunity. Further a number of breakthrough infections have been observed (Bergwerk et al., 2021; Khoury et al., 2021; Naaber et al., 2021; Thomas et al., 2021), suggesting that there are still chances that local clusters happen even in the post-vaccine era.

In the midst of the ongoing pandemic, ways to restart working at offices and teaching at schools in-person while avoiding the risk of transmission have been argued. This is a reasonable argument given the negative impact of online teaching and remote works. A study demonstrated that the shift to online teaching due to the COVID-19 pandemic led to decline in course completion rates among college students (Bird, Castleman, & Lohner, 2020). Further, negative impact of online teaching on mental health, anxiety and depression, has also been suggested (Bird et al., 2020; Huckins et al., 2020; H. Y. Li, Cao, Leung, & Mak, 2020; Sahu, 2020; Wang et al., 2020). For workers, the impact of remote work on productivity is controversial, depending on types of stress (i.e., job related and non-job related) and industries (Bartik, Cullen, Glaeser, Luca, & Stanton, 2020; Baudot & Kelly, 2020; Galanti, Guidetti, Mazzei, Zappalà, & Toscano, 2021; Toscano & Zappalà, 2020). A study demonstrated that the remote work led to static and siloed communication rather than communication between disparate parts, which may hinder employees in gaining new information and connection and lead to low productivity (Yang et al., 2021).

To safely go back to schools and offices, CDC released the guidance to mitigate the risk of infection at schools and offices, including all precautions such as vaccination, universal indoor masking, physical distancing, ventilation, handwashing and respiratory etiquette (Centers for Disease Control and Prevention, 2021a, 2021b). Such guidelines seem to work in limiting the transmission in schools and offices where precautions are implemented (Lessler et al., 2021). Although the precautions implemented in schools and offices limit the risk of transmission within facilities, the risk of transmission outside of facilities cannot be easily controlled, therefore identifying infected and infectious patients as early as possible would further mitigate the risk of transmission in facilities. Indeed, CDC recommend to incorporate screening depending on levels of community transmission and activities that students are involved in (Centers for Disease Control and Prevention, 2021b).

However, the effectiveness of screening in schools and offices is still unclear. Further, it is not well studied which screening strategy effectively identifies infected and infectious individuals to avoid transmission in facilities. For example, we need to choose the type of tests for screening: PCR tests, antigen tests, and antibody tests. The purpose of screening is to identify infected and infectious patients as soon as possible and subsequently performing appropriate treatment and isolation is necessary to mitigate the burden of disease and further transmission. Therefore, fast turnaround time (time between sampling and returning results) and high sensitivity are essential (Larremore et al., 2021). Given that sensitivity is generally higher for PCR tests, whereas the turnaround time is much shorter for antigen tests, there is an argument which test is more appropriate for screening (Larremore et al., 2021). Further screening schedule needs to maximize the effectiveness. In this study, we assess the effectiveness of various screening strategies in schools and offices through simulation and explore the key parameters for effective screening.

## Materials and Methods

### Overview of simulation

The effectiveness of screening is assessed through simulation. For realistic simulation, we develop two models with different scales: a transmission model in a community where the school or office belongs, and a viral dynamics model of each infected individual at the school or the office. The viral dynamics model is a compartment model composed of two components: virus and proportion of uninfected target cell population. The model is parametrized by longitudinal viral load data. The transmission model is also a compartment model composed of four different statuses: susceptible, pre-infectious, infectious, and non-infectious.

Multiple screening strategies were considered. The screening strategies are composed of multiple factors: 1. sensitivity of antigen tests, 2. a screening schedule over the screening period (10 days). For the assessment of effectiveness of screening, we use the proportions of infected cases identified through the screening strategy among all infected cases. Screening simulation was performed 30 times for each setting. Below we describe the detail of the data, the models, and the simulation.

### Viral load data

Longitudinal viral load data of symptomatic and asymptomatic COVID-19 patients were searched through PubMed and Google scholar. The data that meet following criteria are used to estimate parameters of the viral dynamics model: 1) viral loads were measured and reported at least at two time points; 2) samples were collected from upper respiratory specimens, such as nose and pharynx; 3) patients who did not receive antiviral therapy were included. As all the data were extracted from published papers, ethics approval was not required in this study.

### SARS-CoV-2 viral dynamics model and viral load data generation

To describe the temporal change in viral load of each infected individual in the school or the office, a mathematical model describing the viral dynamics of SARS-CoV-2 was employed as in our previous studies (Ejima, Kim, Iwanami, et al., 2021; Ejima, Kim, Ludema, et al., 2021; Iwanami et al., 2021; Jeong et al., 2021; Kim et al., 2021). Briefly explaining, the model is composed of two compartments: the viral load (copies/mL) at time *t,V*(*t*), and the ratio between the number of uninfected cells at time to and that at time 0, *f*(*t*). Note that time 0 is the time of infection, thus *V*(0) = 10 ^−2^ (copies/mL) and *f* (0) = 1.*V*(*t*) draws viral load trajectory which is generally observed in patients infected by virus causing acute infection under reasonable parameter setting: *V*(*t*) initially exponentially increases, then starts declining after hitting the peak. More details are described in **Supplementary Materials**. The model parameters are estimated by fitting the model to the longitudinal viral load data using a nonlinear mixed-effect modelling approach. The nonlinear mixed-effect model allows estimation of population parameters while accounting for variation in parameters between individuals (Best et al., 2017; Gonçalves et al., 2020). As the time of infection is not observed for patients, we estimated the timing of infection as well (Ejima, Kim, Iwanami, et al., 2021; Ejima, Kim, Ludema, et al., 2021). Note that the model parameters were estimated for symptomatic and asymptomatic patients (**Fig. 1C**).

**Figure 1.**
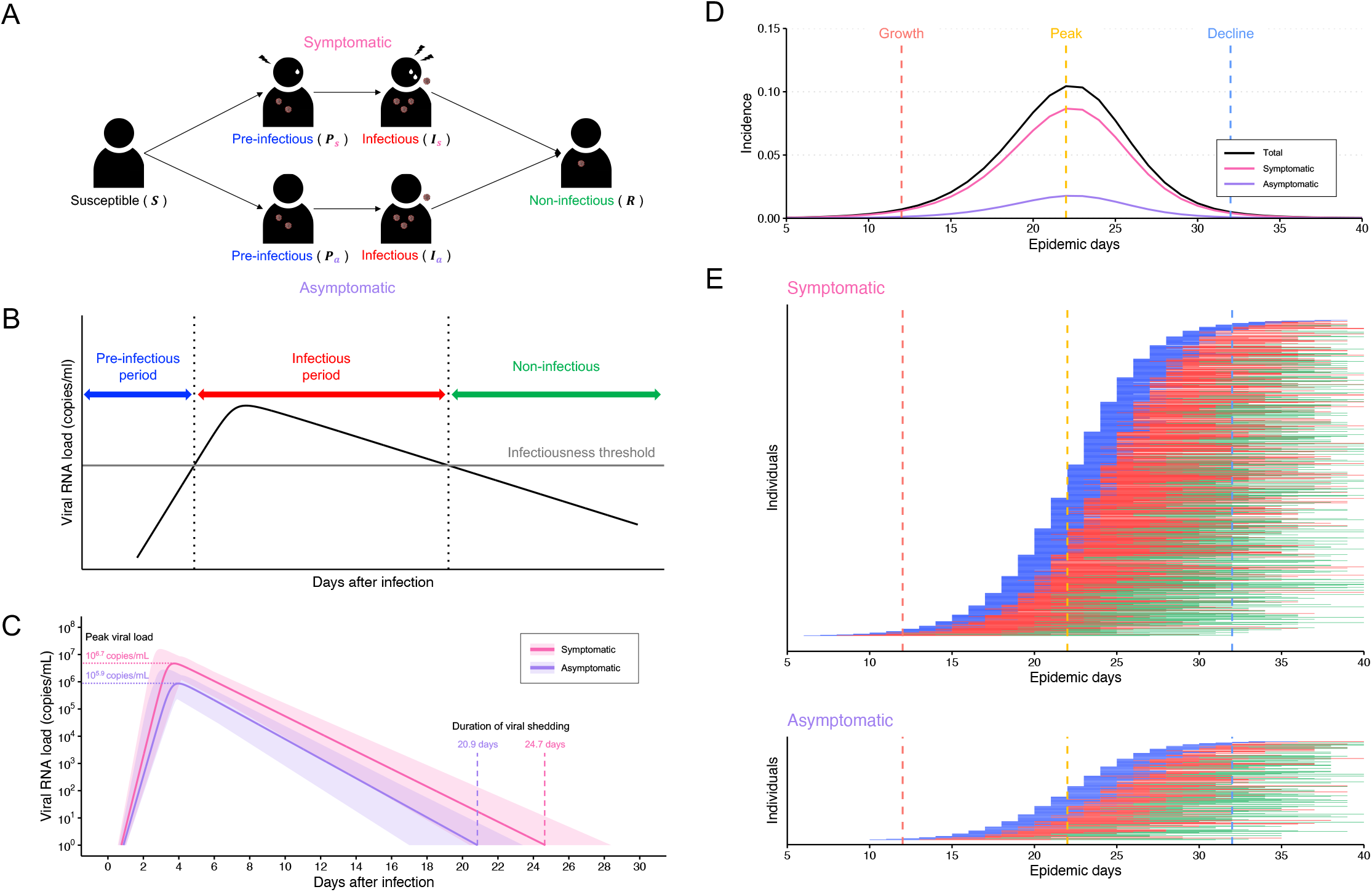
SARS-CoV-2 viral dynamics model and transmission model. **(A)** Schematic illustration of SARS-CoV-2 transmission model. **(B)** Three different phases of SARS-CoV-2 viral dynamics. The black solid line is a typical viral load curve (for an illustration purpose). The horizontal gray solid line is an infectiousness threshold. One can pass three phases since infection: pre-infectious (blue), infectious (red), and non-infectious (green) depending on the viral load, respectively. **(C)** Estimated viral load curves from the SARS-CoV-2 viral dynamics model. The solid lines are drawn using the best fit population parameters (Pink: symptomatic, Purple: asymptomatic). The shaded regions correspond to 95% predictive intervals created using bootstrap approach. **(D)** SARS-CoV-2 epidemic curves. The black, pink, and purple curves are the daily incidence (proportion to the total population) of new infections, symptomatic, and asymptomatic infections, respectively. The vertical red, yellow, and blue dashed lines are the timings of screening initiation for the growth, peak, and decline phases of epidemic, respectively. **(E)** Simulated SARS-CoV-2 viral dynamics of symptomatic and asymptomatic individuals in the school or office over time. The blue, red, and green lines are pre-infectious, infectious, and non-infectious stages of each case as defined in **(B)**. Note that green line disappears when the viral load drops below detection limit of 10^2^ copies/mL.

The estimated parameters are used for simulating viral load trajectories of those infected cases in the school or office. To account for the measurement error, we also computed “measured” viral load, 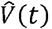, in addition to “true” viral load, *V*(*t*) (Jeong et al., 2021). Specifically, longitudinal true viral load for a patient *k, V*_*k*_(*t*), is generated by running the viral dynamics model with a parameter set, which is resampled from the estimated distributions. The measured viral load for the patient *k* is computed by adding an error term: 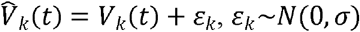. The variance of the error term, *σ* is estimated by fitting normal distribution to the residuals (i.e., the difference between the estimated viral load and measured viral load) obtained in the model fitting.

### SARS-CoV-2 transmission model

We further developed a mathematical model to describes the epidemiological dynamics of COVID-19 in a community which the school or office belongs. The model is an extension of the so-called SIR model, composed of four different groups divided by infectiousness and immune status: Susceptible (*S*), Pre-infectious (*P*), Infectious (*I*), and Non-infectious (and immune) (*R*) (**Fig. 1A**). We further divided the *P* and *I* compartments by the presence of symptoms (*p*: the proportion of not presenting symptoms). Full details of the transmission model are provided in **Supplementary Materials**.

The parameters related to the disease progression are informed from the viral dynamics model (**Fig. 1B**). Specifically, the pre-infectious period, 1/*ε*, is defined as the time since infection to the time when the viral load passes the infectiousness threshold (=10^5^ copies/mL (Goyal, Reeves, Cardozo-Ojeda, Schiffer, & Mayer, 2021; Marc et al., 2021; van Kampen et al., 2021; Wölfel et al., 2020)), and the infectious period, 1*/σ*, is defined as the time during which viral load is above the infectiousness threshold. We estimated the values of these parameters by running the viral dynamics model with 1000 parameter sets resampled from the estimated distributions. The median values from the distributions were used for 1/*ε* and 1/*σ*. Note that this process is performed for symptomatic cases and asymptomatic cases independently.

The proportion of asymptomatic cases, *p*, is set as 0.17 (Byambasuren et al., 2020). The transmission rate, *b* is assumed constant over the infectious period and defined using the basic reproduction number, *R*_0_, and other parameters as follows: 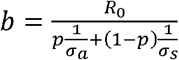 (the derivation process is available in **Supplementary Materials**), where 1/*σ*_*s*_ and 1/*σ*_*a*_ are infectious periods for symptomatic and asymptomatic cases. *R*_0_ is set as 7.0, which is an estimated value for the B.1.617.2 (delta) variant (Burki, 2021). All estimated and assumed model parameters are summarized in **Table S3**.

### Simulation of screening COVID-19 patients

Using the above two models (i.e., the viral dynamics model and the transmission model), we assess the effectiveness of various screening strategies. The effectiveness is assessed by three metrics counting infected cases: the proportions of infected cases (pre-infectious, infectious, and non-infectious) identified through the screening strategy among all infected cases. We further computed the breakdown as we believe that identifying pre-infectious and infectious patients and identifying non-infectious patients have different implications from public health perspectives. We computed these metrics for those infected (a) at the start of screening and (b) at the start of screening and during the screening period (10 days). In other words, the latter includes those infected during the screening period (thus in addition to the numerator [identified cases], the denominator [all infected cases] increases during the screening period).

The size of the school or the office is assumed to be 1000. Their risk of infection is determined by the risk of community, thus if the total attack rate in the community by the end of the epidemic is *AR*, then final size of the school or the office is 1000 * *AR. AR* is derived from the final size equation (1 − *AR* exp (−*R*_0_ * *AR*)). The timing of infection is determined following the distribution of epidemic curve. Among all cases, 1000 * *AR* * (1 − *p*) cases develop symptoms.

Several different scenarios varying the sensitivity of antigen tests and the schedule of screening during the screening period (10 days) are considered. The turnaround time is assumed negligible for antigen tests (Larremore et al., 2021). The detection limits for high sensitivity antigen tests and low sensitivity antigen tests are 6.3 × 10^4^ copies/mL and 2.0 × 10^6^ copies/mL (Yamaoka et al., 2021), respectively.

Screening with antigen tests is performed four times with different schedules over the screening period (10 days): (1) everyday (Day 0, 1, 2, 3), (2) two days interval (Day 0, 2, 4, 6), (3) three days interval (Day 0, 3, 6, 9), (4) late screening (Day 0, 8, 9, 10), and (5) random screening (the timing of test is determined randomly over the 10 days) (**Fig. 2A**). We also performed the simulation with daily screening to explore key parameters associated with high effectiveness of the screening. Beside the screening tests, patents are further identified through symptom presence. Thus symptomatic cases could be identified either symptom onset (an incubation period of each case is randomly determined from the distribution previously estimated: *lognorm*(1.76, 0.41) [the median incubation period is 5.8 days] (Ejima, Kim, Ludema, et al., 2021)) or positive screening test results that come first.

**Figure 2.**
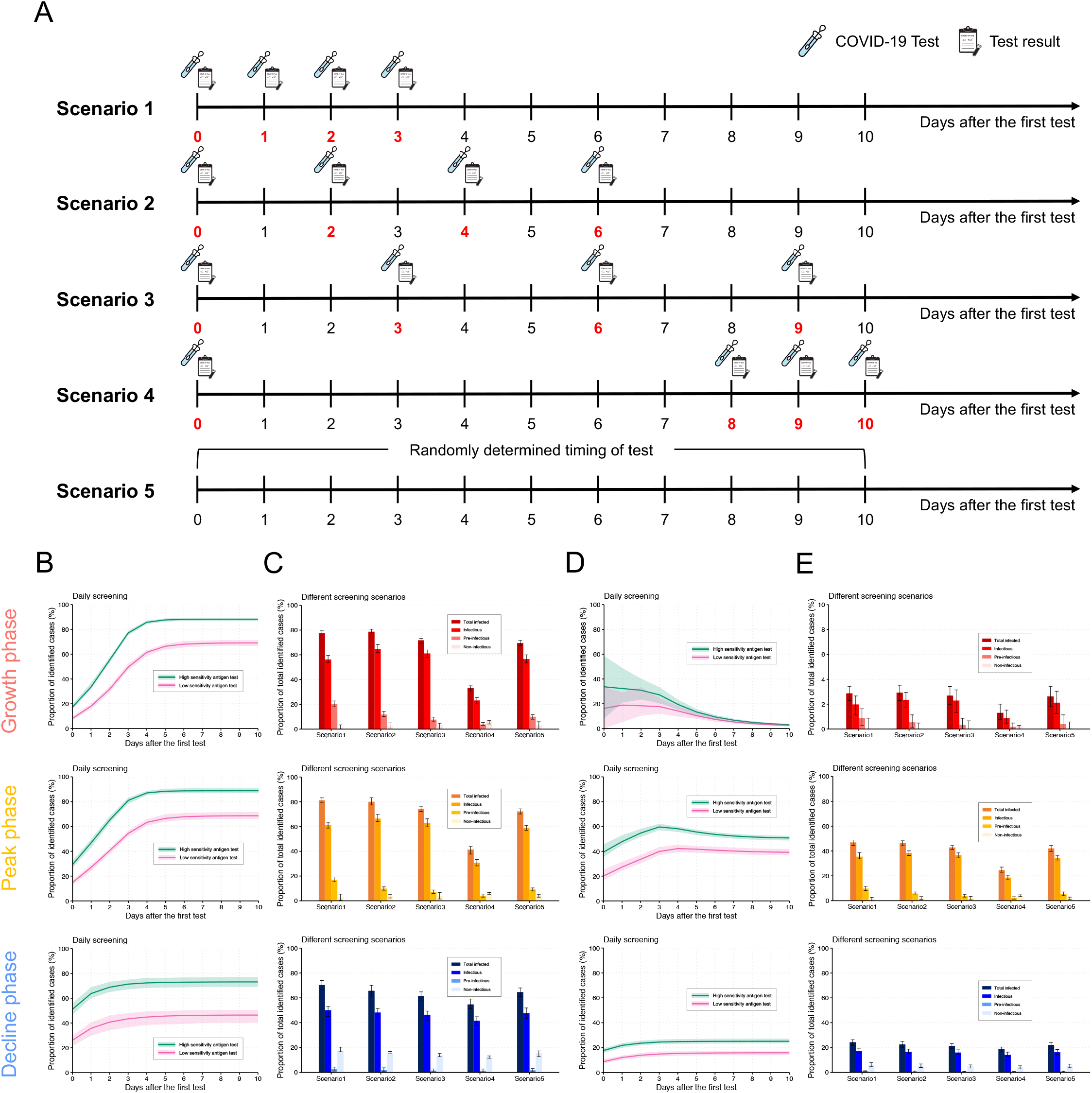
Effectiveness of screening strategies without/with considering cases infected during screening period. **(A)** Different screening scenarios with high sensitivity antigen tests. **(B)** Cumulative proportion of identified cases by daily screening with low (pink) and high (green) sensitivity antigen tests without considering cases infected during screening. The shaded regions are 95% CIs of 30 independent simulations. **(C)** Proportion of identified cases under different screening scenarios for high sensitivity antigen tests without considering cases infected during screening. In addition to total infected cases, cases infectious, pre-infectious, and non-infectious when they are identified are separately counted. The error bars are 95% CIs of 30 independent simulations. **(D)** Cumulative proportion of identified cases by daily screening with low (pink) and high (green) sensitivity antigen tests with considering cases infected during screening. The shaded regions are 95% CIs of 30 independent simulations. **(E)** Proportion of total identified cases under different screening scenarios for high sensitivity antigen tests with considering cases infected during screening. Note that in **(B)**-**(E)**, first, second, and third rows of panels correspond to the results for the screening initiated in the growth, peak, and decline phases of the epidemic, respectively.

Screening is initiated at different epidemic phases: growth, peak, and declining phases. Note that the epidemiological dynamics would not be influenced by screening tests, because we assume a community of large population and a school or an office of considerably small population. Further, we assume that no transmission occurs in the school or the office, which is a reasonable assumption given the observation that the transmission risk is limited at schools and offices where precautions are implemented (Lessler et al., 2021). Therefore, we assumed identified cases at the school or office are infected in the community, but outside of the facility. In other words, the risk of infection for those in the school or the office is the same as the risk in the community. As sensitivity analyses, the simulation was performed varying the number of antigen tests (2 to 5) and the length of the screening period (10 to 30 days).

## Results

### Different virus infection dynamics between symptomatic and asymptomatic patients

10 papers were identified which met all the inclusion criteria. In total, the viral load data of 109 symptomatic patients and 101 asymptomatic patients were used for parameter estimation of the viral dynamics model. Seven studies were from Asia and two were from USA. The other was from Europe (**Table S1**).

The viral dynamics model was fitted to the longitudinal viral load data from the symptomatic and asymptomatic patients (**Fig. S1, S2** and **Table S2**). We observed difference on the peak viral load and the duration of viral shedding between groups, but not on the time of the peak viral load (**Fig. 1C**). Considering viral load dynamics observed here, we evaluated effectiveness of screening strategies as below.

### Effectiveness of screening strategies

The daily incidence at the community peaks at the epidemic day 22 and the epidemic disappears in 40 days because of the high transmission potential of the delta variant (*R*_0_ *=* 7.0) (**Fig. 1D**), thus *AR* ≅ 1.0. For the screening simulation, the screening starts at epidemic day 12, 22, and 32, each of which corresponds to growth, peak, and decline phases of the epidemic. Because the infection risk for those at the school and the office is dependent on that of the community, the epidemic curve at the facility is proportional to that of the community (**Fig. 1E**). At the growth phase, more cases at the first screening are pre-infectious and with low viral load (24%) because time does not pass much since infection by the time, whereas more cases are already infectious and with high viral load at the decline phase (42%) (**Fig. S3**).

The effectiveness of different screening strategies was assessed based on the proportion of identified cases through the screening. To identify key parameters associated with high effectiveness of screening, we first performed simulation with daily screening. When those who infected during the screening period were not included in the effectiveness assessment, the proportion of identified cases monotonically increases during the screening period, as they are identified by multiple testing or symptom presence (**Fig. 2B and Fig. S4A**). The proportion saturates around Day 5 because the viral load drops below the detection limit of the tests for most cases who infected before the initiation of screening by this time. Screening using high sensitivity antigen tests could identify twice as many as patients compared with that using low sensitivity antigen tests on Day 0, and the effectiveness remains higher over the screening period.

The temporal behavior of the proportion of identified cases is different depending on the epidemic phase when the screening is performed. At the growth phase, the first screening test cannot capture the cases who have not presented symptoms (but will present symptoms later) and those with low viral load below the detection limit because more cases are recently infected (**Fig. S3**), thus the proportion is lower than that for the decline phase: 8% (95%CI: 6-10) and 17% (95%CI: 15-19) for low and high sensitivity antigen tests, respectively (**Fig. 2B top**). The proportion reaches 69% (95%CI: 66-71) and 88% (95%CI: 86-89) by the end of screening (**Fig. 2B top**). When the screening starts at the decline phase, more patents could be identified by the first day because of symptom presence and high viral load (**Fig. 2B bottom**); however, the proportion reaches 46% (95%CI: 40-50) and 73% (95%CI: 69-77) by the end of screening with low and high sensitivity antigen tests, respectively (**Fig. 2B bottom**). The proportions with the screening at the decline phase cannot reach those with the screening at the growth phase, because more patients are already non-infectious and the viral load drops below the detection limits (**Fig. S3**). Note that symptomatic patients were detected by either screening tests or symptom presence (**Fig. S4A**). Overall, high sensitivity antigen tests can identify about 20% more cases under everyday screening regardless of the timing of screening initiation.

The simulation is performed for different scenarios with high sensitivity antigen tests due to its high effectiveness (**Fig. 2A**). We primarily focus on the proportion of cases identified through simulation, and further computed the breakdown (i.e., pre-infectious, infectious, and non-infectious cases). Regardless of the epidemic phase, early and frequent screening (such as Scenarios 1 and 2, which perform test every day or with two days interval) can identify more cases because cases are identified when their viral load is still high (**Fig. 2C**). Indeed, when the screening is started at the growth phase, Scenario 1 (everyday screening) yielded to 77% (95% CI: 75-79) of total cases identified, whereas it was 33% (95% CI: 31-35) for Scenario 4 (late screening). The difference between scenarios shrinks when the screening is performed at the decline phase due to limited new infections. Such difference becomes clear when we focus on the proportion of identified pre-infectious cases. The contribution of pre-infectious to the proportion of total cases is substantial when the screening is performed at the growth phase (20% under Scenario 1), whereas more cases are identified at non-infectious phase when the screening is performed at the decline phase (18% under Scenario 1). We also found that the proportion of identified asymptomatic cases are influenced by scenarios and epidemic phases (see **Fig. S4B**). Over 90% of asymptomatic patients are identified by early and frequent screening (Scenario 2) in the growth phase, but the proportion drops to around 60% even under the best scenario (i.e., Scenario 1) in the decline phase because many of asymptomatic patients are infected long before the screening and their viral load is below the detection limit at the timing of screening.

Sensitivity analyses were performed by varying the number of high sensitivity antigen tests and the length of screening period to see if the best scenario (with the highest proportion of identified cases) could be influenced by those parameters (**Fig. S4C**). The length of the screening period did not impact on the best scenario; however, the best scenario shift towards less frequent testswhen the number of antigen tests is limited especially when the screening is performed at the growth phase. This is because some of pre-infectious cases under detection limit cannot be identified at the first few days; in other words, the best scenario was robust when the screening is performed at the decline phase due to the limited number of new infection (thus pre-infectious cases).

We further performed the analyses including those infected during the screening period (**Fig. 2DE** and **Fig. S5**). Compared with the analyses excluding those infected during the screening period, as both the numbers of cases (denominator) and identified cases (numerator) increase over time, the proportion of identified cases does not necessarily increase, which is more drastic when the screening is performed when epidemic is still growing (**Fig. 2D top**). Under the 5 scenarios for the growth phase, fewer than 4% of total infected individuals are identified (**Fig. 2E top**). In the peak phase, about 50% of infected cases are identified by the daily screening (**Fig. 2D middle**). This is because both the limited new incidence and viral loads which are high enough to be identified (**Fig. 1E and S3**). Under the 5 scenarios, Scenario 1 and 2 identified around 50% of infected cases (**Fig. 2E middle**). Among those identified cases by screening at the peak phase, 60% of asymptomatic patients and 40% of pre-symptomatic patients were identified by early and frequent screening (Scenario 1 and 2), respectively (**Fig. S5B, middle**). As new cases are limited when the screening is performed at the decline phase, the proportion looks monotonically increasing (**Fig. 2D bottom**). In this phase, the high sensitivity antigen tests identified about 20% of infected cases regardless of the scenarios (**Fig. 2E, bottom**). Other results such as the best scenario were consistent with those when only cases infected before the screening initiation were considered (**Fig. S5C**).

## Discussion

Our society is seeking for approaches to safely restart economic and educational activities involving in-person interactions in midst of the pandemic. Along with precautious measures, screening is considered as a key to mitigate the risk of infection allowing in-person interactions. We assessed the effectiveness of screening strategies using antigen tests through simulation.

In the simulation, the transmission model and the viral dynamics model were combined to incorporate both epidemiological dynamics and within-person biological dynamics, because both factors influence the assessment of the effectiveness of screening strategies. To parameterize the model, we used longitudinal viral load data for the viral dynamics model and epidemiological parameters, a part of which were informed from the viral dynamics model, for the transmission model. To identify key parameters for effective strategies, we first performed daily screening initiated at different epidemic phases (early, peak, decline phases). If the screening is performed every day for 10 days, 69 % and 88 % of infected individuals could be identified by screening with low and high sensitivity antigen tests, respectively. The proportion declines to 46 % and 73 % when the screening is initiated at the decline phase of the epidemic; however, this is mainly because of those cases who are already not infectious. Therefore, high sensitivity antigen tests yielded 20% more cases identified through the everyday screening regardless of the timing of screening initiation.

Next, we further performed simulation to assess the effectiveness of screening under different schedules, varying four times tests (high sensitivity antigen tests) over 10 days of the screening period. Regardless of the epidemic phase, we found early and frequent screening yielded high effectiveness, because more cases are identified while their viral load is high. Sensitivity analyses revealed that the best effective scenario shifts toward less frequent tests when the number of antigen tests is limited especially when the screening is performed at the growth phase. Overall, our simulation suggested early and frequent screening with high sensitivity antigen tests can identify more cases.

This is the first study that assessed the effectiveness of screening strategies with antigen tests in schools or offices. There is a study focusing on entire population, suggesting that frequent screening tests with fast turnaround time is key to mitigate the transmission risk in the population (Larremore et al., 2021). Compared with this study, our study is unique because we focused on a small group (i.e., a school or an office) as the screening target, given that screening in a huge population does not seem realistic. Therefore, the screening does not influence on the epidemiological dynamics in the community, and we could separately argue the transmission in the community and the screening effectiveness in a school or an office. Further we assumed the limited screening period. Although antigen screening is less expensive compared with PCR tests, enforcing all people to get tested over the pandemic period is challenging.

In this study, we used antigen tests rather than PCR testing, because using PCR testing for screening purpose is practically challenging given the cost and turnaround time (2 days (Larremore et al., 2021)). There are multiple antigen tests with varying sensitivity (Yamaoka et al., 2021). Although our study did not investigate the difference in cost between different antigen tests, we found that high sensitivity antigen tests among all antigen tests available so far yielded 20% more identified cases than low sensitivity antigen tests if the screening is implemented every day over 10 days.

We did not directly consider the cost of antigen tests, however, if available antigen tests are limited, early and frequent screening might not be the best strategy. Rather than that, screening with a few days’ interval might be an option to wait until those who have just been infected with low viral load will be detected by antigen tests. Under resource limited circumstances, the screening schedule needs to be reconsidered. However, our computational framework is flexible enough to assess the effectiveness of screening strategy accounting for such situation.

Regardless of the flexibility of the simulation framework, there are a few limitations which need to be addressed in future studies. First, we did not consider false-positive rate of the antigen tests (specificity of antigen tests for SARS-CoV-2: 99.6% (Dinnes et al., 2021)). Especially in screening settings under low prevalence, even a small false positive rate would be an issue. For example, if the prevalence is 1% (and the sensitivity is 100%), the screening test with 99% sensitivity will produce almost the same number of false negative cases. As people with false positive results may need to be confirmed by PCR tests, targeted screening (targeting high prevalence population) should be considered. Second, both the transmission model and the viral dynamics model did not consider emerging variants, reinfection, breakthrough infection, and vaccine effect. If the viral dynamics of delta variant is different from the original variant (Li et al. suggested that the delta variant presented faster viral replication (B. Li et al., 2021)), our model needs to be updated. However, we anticipate that faster viral replication would not influence our findings because the replication speed is associated with the length that the virus cannot be detected, which is already less than 3 days with the original variant. The models need to update the transmission risk of under vaccination. Third, the transmission model did not incorporate behavioral change during the pandemic or screening. The contact pattern dynamically changed as a response to the pandemic, which yielded multiple waves observed in most of the countries (Crane, Shermock, Omer, & Romley, 2021; Feehan & Mahmud, 2021; Zhang et al., 2020), thus such behavioral dynamics needs to be incorporated into the transmission model.

In conclusion, to safely open our economic and educational activities, we recommend that frequent and early screening with high sensitivity antigen tests will substantially mitigate the risk of transmission in a school or an office. We believe that our computational framework would be helpful to assess the effectiveness of various screening strategies under different circumstances including emergence of new variants.

## Data Availability

All data produced in the present study are available upon reasonable request to the authors

## Acknowledgments

We would like to thank Yoshihiro Okada, Junichi Katada, Atsuhiko Wada, Kaku Irisawa and Toshiki Takei at FUJIFILM Corporation for useful discussion. This study was supported in part by Basic Science Research Program through the National Research Foundation of Korea funded by the Ministry of Education 2019R1A6A3A12031316 (to K.S.K.); Grants-in-Aid for JSPS Scientific Research (KAKENHI) B 18KT0018 (to S.Iwami.), 18H01139 (to S.Iwami.), 16H04845 (to S.Iwami.), Scientific Research in Innovative Areas 20H05042 (to S.Iwami.); AMED Strategic International Brain Science Research Promotion Program (to K.A.); AMED CREST 19gm1310002 (to S.Iwami.); AMED Japan Program for Infectious Diseases Research and Infrastructure, 20wm0325007h0001 (to S.Iwami.), 20wm0325004s0201 (to S.Iwami.), 20wm0325012s0301 (to S.Iwami.), 20wm0325015s0301 (to S.Iwami.); AMED Research Program on HIV/AIDS 19fk0410023s0101 (to S.Iwami.); AMED Research Program on Emerging and Re-emerging Infectious Diseases 19fk0108050h0003 (to S.Iwami.), 19fk0108156h0001 (to S.Iwmai.), 20fk0108140s0801 (to S.Iwami.) and 20fk0108413s0301 (to S.Iwami.); AMED Program for Basic and Clinical Research on Hepatitis 19fk0210036h0502 (to S.Iwami.); AMED Program on the Innovative Development and the Application of New Drugs for Hepatitis B 19fk0310114h0103 (to S.Iwami.); JST MIRAI (to S.Iwami.); Moonshot R&D Grant Number JPMJMS2021 (to K.A. and S.Iwami.) and JPMJMS2025 (to S.Iwami.); Mitsui Life Social Welfare Foundation (to S.Iwami.); Shin-Nihon of Advanced Medical Research (to S.Iwami.); Suzuken Memorial Foundation (to S.Iwami.); Life Science Foundation of Japan (to S.Iwami.); SECOM Science and Technology Foundation (to S.Iwami.); The Japan Prize Foundation (to S.Iwami.); Daiwa Securities Health Foundation (to S.Iwami.); the MIDAS Coordination Center (MIDASSUGP2020-6) by a grant from the National Institute of General Medical Science (3U24GM132013-02S2) (to K.E.). The study does not necessarily represent the views of the funding agencies listed above.

## AUTHOR CONTRIBUTIONS

KE, SIwami and KA designed the research. YDJ, KE, KSK, SI and SIwami carried out the computational analysis. KE and SIwami supervised the project. All authors contributed to writing the manuscript.

