## Supplementary Information for "Safely return to schools and offices: early and frequent screening with high sensitivity antigen tests effectively identifies COVID-19 patients"

^1^interdisciplinary Biology Laboratory (iBLab), Division of Biological Science, Graduate School of Science, Nagoya University, Nagoya, Japan. ^2^Department of Mathematics, Pusan National University, Busan, South Korea. ^3^Department of Epidemiology and Biostatistics, Indiana University School of Public Health-Bloomington, IN, USA. ^4^Institute of Mathematics for Industry, Kyushu University, Fukuoka, Japan. ^5^Institute for the Advanced Study of Human Biology (ASHBi), Kyoto University, Kyoto, Japan. ^6^NEXT-Ganken Program, Japanese Foundation for Cancer Research (JFCR), Tokyo, Japan. ^7^Science Groove Inc., Fukuoka, Japan. ^8^International Research Center for Neurointelligence, The University of Tokyo, Tokyo, Japan.

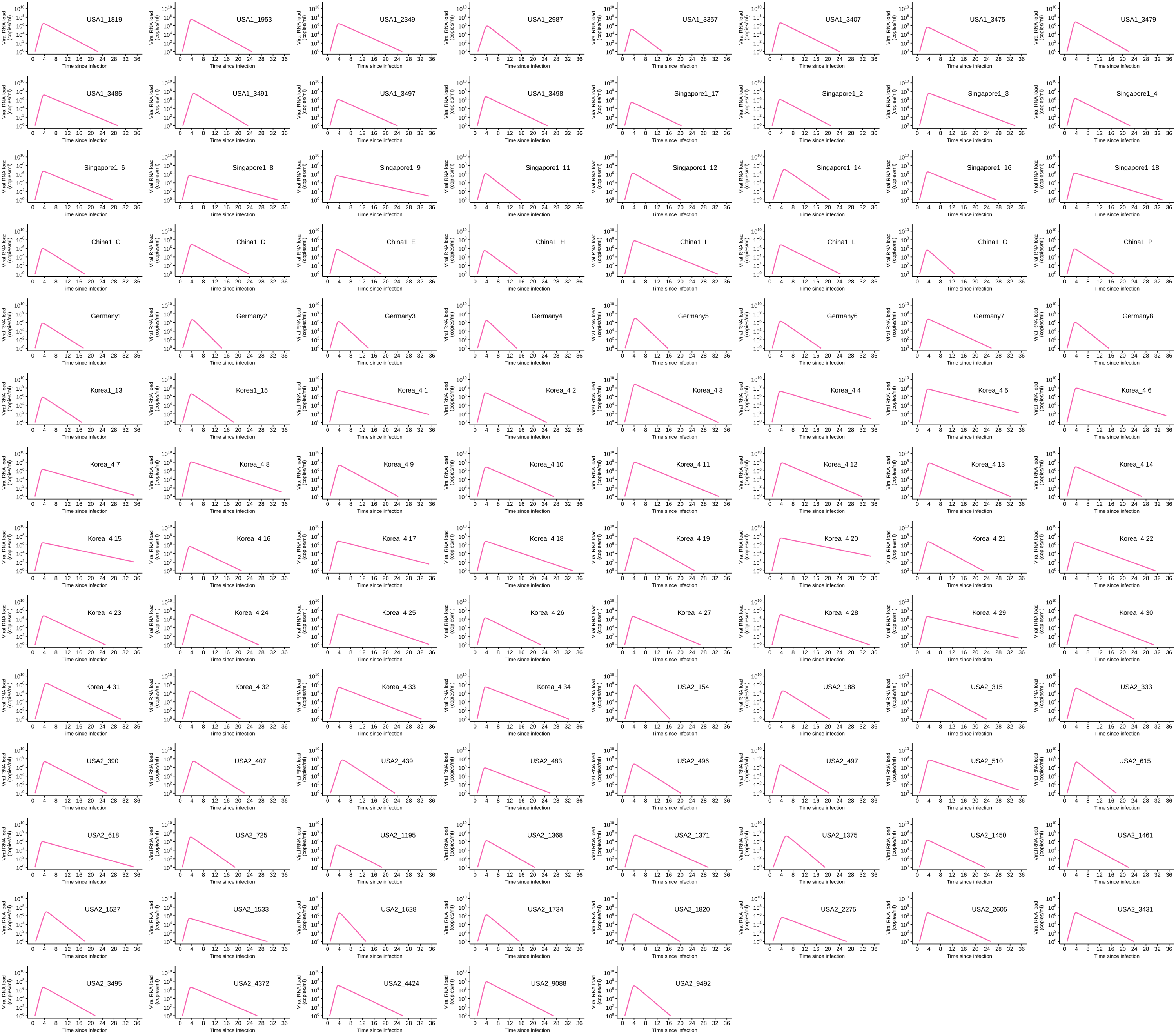

**Figure S1.** **Viral load trajectory for 109 symptomatic patients.** The estimated viral load for each individual (solid lines) along with the observed data (closed dots) are depicted using the best-fit parameter estimates.

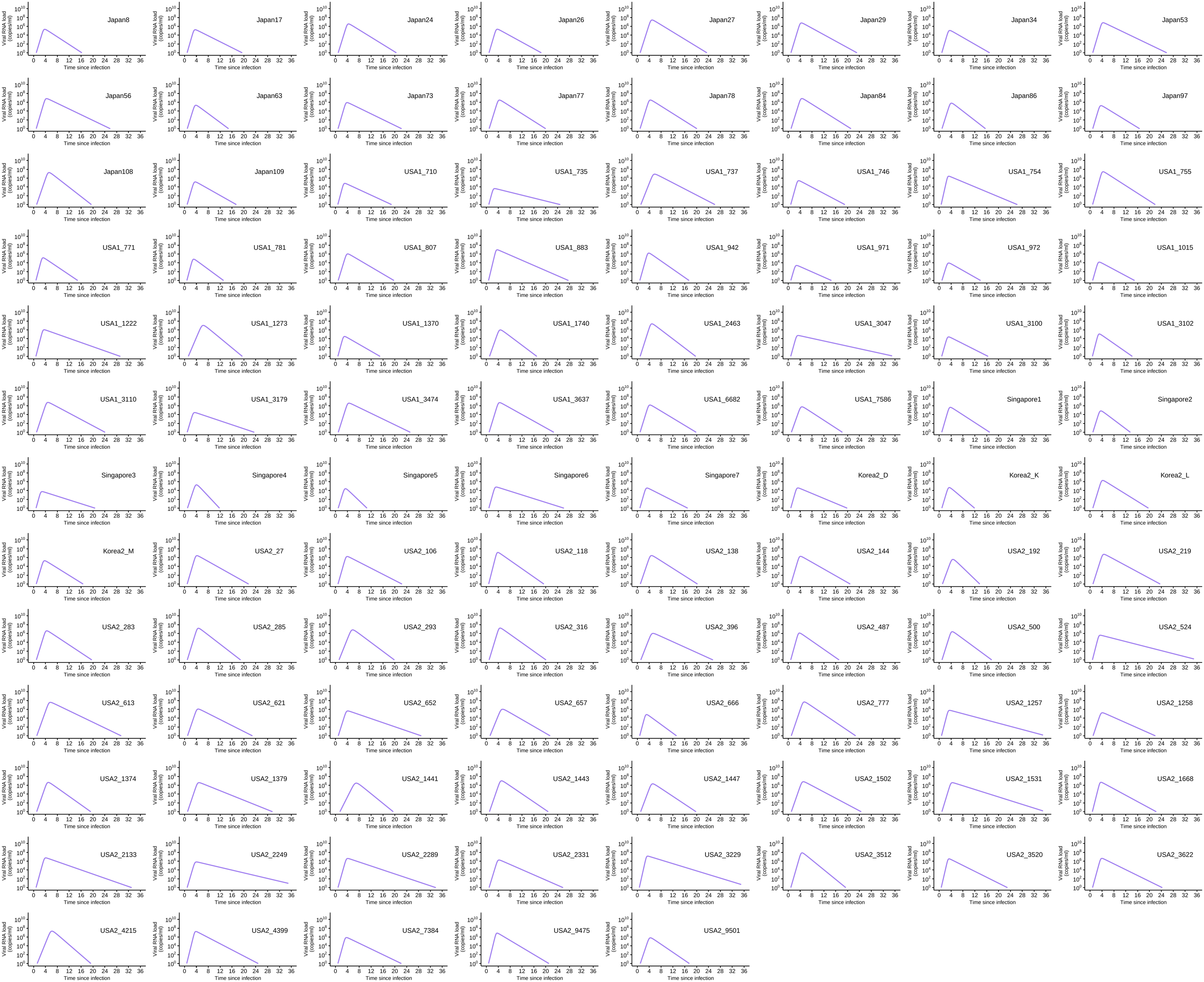

**Figure S2. Viral load trajectory for 101 asymptomatic patients.** The estimated viral load for each individual (solid lines) along with the observed data (closed dots) are depicted using the best-fit parameter estimates.

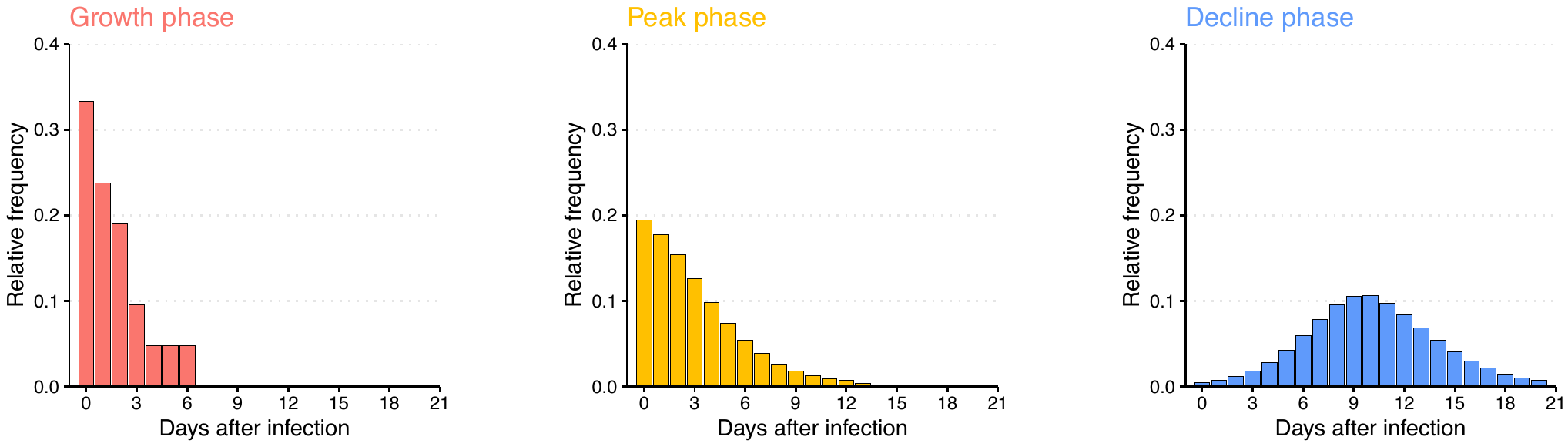

**Figure S3. Distribution of days after infection at each epidemic phase.** The red, orange, and blue bars correspond to the growth, peak, and decline phases, respectively.

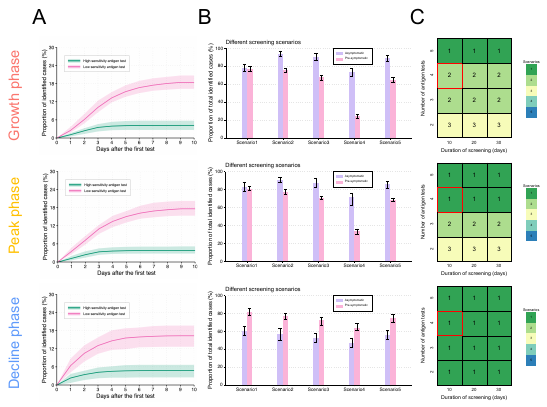

**Figure S4. Effectiveness of screening without considering cases infected during screening.** **(A)** Cumulative proportion of identified cases by the presence of symptoms (not by antigen tests). The pink and green lines indicate the scenarios with low and high sensitivity antigen tests, respectively. The shaded regions are 95%CIs of 30 independent simulations. **(B)** Proportion of identified asymptomatic (purple) and pre-symptomatic (pink) cases under different screening scenarios for high sensitivity antigen test. The error bars are 95%CIs of 30 independent simulations. **(C)** Best screening scenarios under different numbers of high sensitivity antigen tests and durations of screening. The red square corresponds to the baseline setting of screening (i.e., four times of tests over 10 days). Note that in **(A)**-**(C)**, first, second, and third rows of panels correspond to the growth, peak, and decline phases of the epidemic, respectively.

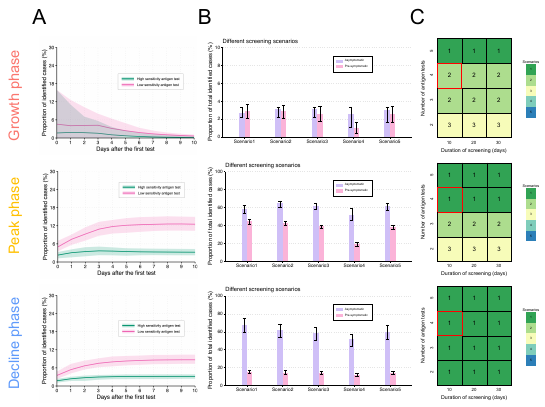

**Figure S5. Effectiveness of screening with considering cases infected during screening.** **(A)** Cumulative proportion of identified cases by the presence of symptoms (not by antigen tests). The pink and green lines indicate the scenarios with low and high sensitivity antigen tests, respectively. The shaded regions are 95%CIs of 30 independent simulations. **(B)** Proportion of identified asymptomatic (purple) and pre-symptomatic (pink) cases under different screening scenarios for high sensitivity antigen test. The error bars are 95%CIs of 30 independent simulations. **(C)** Best screening scenarios under different numbers of high sensitivity antigen tests and durations of screening. The red square corresponds to the baseline setting of screening (i.e., four times of tests over 10 days). Note that in **(A)**-**(C)**, first, second, and third rows of panels correspond to the growth, peak, and decline phases of the epidemic, respectively.

**Table S1. Summary of the viral load data**

| **Country** | **Number of cases** | **Reporting unit** | **Specimens for measuring viral load** | **Source** |
| --- | --- | --- | --- | --- |
| **Symptomatic** |  |  |  |  |
| USA | $33$ | cycle threshold^#^ | Nares and oropharyngeal swabs | (Kissler et al., 2021) |
| USA | $12$ | cycle threshold^#^ | Nares and oropharyngeal swabs | (Kissler et al., 2020) |
| Germany | $8$ | viral load (copies/swab)^&^ | Pharyngeal swab | (Wölfel et al., 2020) |
| Korea | $34$ | cycle threshold^#^ | Oro/nasopharyngeal swabs | (Jang, Rhee, Wi, & Jung, 2021) |
| Korea | $2$ | cycle threshold^#^ | Oro/nasopharyngeal swab | (E. S. Kim et al., 2020) |
| Singapore | $12$ | cycle threshold^#^ | Nasopharyngeal swab | (Young et al., 2020) |
| China | $8$ | cycle threshold^#^ | Nasal swab | (Zou et al., 2020) |
| **Asymptomatic** |  |  |  |  |
| USA | $44$ | cycle threshold^#^ | Nares and oropharyngeal swabs | (Kissler et al., 2021) |
| USA | $28$ | cycle threshold^#^ | Nares and oropharyngeal swab | (Kissler et al., 2020) |
| Japan | $18$ | cycle threshold^#^ | Nasopharyngeal or throat swab | (Sakurai et al., 2020) |
| Korea | $4$ | cycle threshold^#^ | Nasal and throat swabs | (S. E. Kim et al., 2020) |
| Singapore | $7$ | cycle threshold^#^ | Nasopharyngeal swab | (Kam et al., 2021) |

^#^Viral load was calculated from cycle threshold values using the conversion formula: $\log_{10} \left( Viral laod \left[ copies/mL \right] \right)=-0.32\times Ct values \left[ \mathrm{cycles} \right]+14.11$

(Peiris et al., 2003)

^&^1 swab = 3 mL (Wölfel et al., 2020).

**Table S2. Estimated parameters for the SARS-CoV-2 viral dynamics model**

| **Parameters** | **Symbol** | **Unit** | **Symptomatic** | **Asymptomatic** |
| --- | --- | --- | --- | --- |
| Maximum rate constant for viral replication | $\gamma$ | day^-1^ | $6.81$ | $6.06$ |
| Rate constant for virus infection | $\beta$ | (copies/mL)^-1^ day^-1^ | $9.41\times{10}^{-7}$ | $4.15\times{10}^{-6}$ |
| Death rate of infected cells | $\delta$ | day^-1^ | $0.74$ | $0.83$ |

**Table S3. Estimated or assumed parameters for SARS-CoV-2 transmission model**

| **Parameters** | **Symbol** | **Unit** | **Value** | **Source** |
| --- | --- | --- | --- | --- |
| Basic reproduction number | $R_{0}$ | $-$ | $7$ | (Burki, 2021) |
| Transmission rate | $b$ | day^-1^ | $1.1$ | Theoretically derived^$^ |
| Proportion of not presenting symptoms | $p$ | $-$ | $0.17$ | (Byambasuren et al., 2020) |
| Pre-infectious period for symptomatic individuals | $1/\epsilon_{s}$ | day | $2.7$ | Estimated^*^ |
| Pre-infectious period for asymptomatic individuals | $1/\epsilon_{a}$ | day | $3.2$ | Estimated^*^ |
| Infectious period for symptomatic individuals | $1/\sigma_{s}$ | day | $6.6$ | Estimated^*^ |
| Infectious period for asymptomatic individuals | $1/\sigma_{a}$ | day | $5.3$ | Estimated^*^ |

$ $b=\frac{R_{0}}{p\frac{1}{\sigma_{a}}+\left( 1-p \right)\frac{1}{\sigma_{s}}}$ * The median value was used for the simulation.

**Supplementary Note 1: SARS-CoV-2 viral dynamics model**

We used a mathematical model of SARS-CoV-2 viral dynamics without antiviral treatment previously proposed in (Ejima, Kim, Iwanami, et al., 2021; Ejima, Kim, Ludema, et al., 2021; Iwanami et al., 2021; Jeong et al., 2021; Kim et al., 2021):

$$\frac{df(t)}{dt}=-\beta f\left( t \right)V\left( t \right),$$

$$\frac{dV(t)}{dt}=\gamma f\left( t \right)V\left( t \right)-\delta V\left( t \right),$$

where $f\left( t \right)$ is the ratio between the number of uninfected target cells at time $t$ and the number of uninfected target cells at the infection time, and $V(t)$ is the amount of virus per unit of sample specimens (copies/mL) at time $t$. The parameters $\gamma$, $\beta$, and $\delta$ represent the maximum viral replication rate, the rate constant for virus infection, and the death rate of infected cells, respectively.

**Supplementary Note 2: SARS-CoV-2 transmission model**

We developed a deterministic compartment model for SARS-CoV-2 transmission in a community. The model is composed of four different groups divided by infectiousness and immune status: the proportions of susceptible, $S\left( t \right),$ pre-infectious, $P\left( t \right)$, infectious, $I\left( t \right)$, and non-infectious (and immune), $R\left( t \right)$ (**Fig. 1C**). Accounting for presence and absence of symptoms, $P\left( t \right)$ and $I\left( t \right)$ are separated into two groups (the subscripts $a$ and $s$ correspond to symptomatic and asymptomatic, respectively). Newly infected cases are immediately allocated to either $P_{a}$ or $P_{s}$ following the proportion of not presenting symptoms, $p$. Those in $P\left( t \right)$move to $I\left( t \right)$ once they acquire infectiousness, which corresponds to the timing when the viral load reaches the infectiousness threshold (${10}^{5}$ copies/mL) (**Fig. 1B**). Those in $I\left( t \right)$ move to $R\left( t \right)$ once they lose infectiousness, which corresponds to the timing when the viral load drops below the infectiousness threshold. The model can be described by following ordinary differential equations:

$$\frac{dS(t)}{dt}=-bS\left( t \right)(I_{a}\left( t \right)+I_{s}\left( t \right)) ,$$

$$\frac{dP_{a}(t)}{dt}=pbS\left( t \right)(I_{a}\left( t \right)+I_{s}\left( t \right))-\varepsilon_{a}P_{a}(t),$$

$$\frac{dP_{s}(t)}{dt}=(1-p)bS\left( t \right)(I_{a}\left( t \right)+I_{s}\left( t \right))-\varepsilon_{s}P_{s}(t),$$

$$\frac{dI_{a}(t)}{dt}=\varepsilon_{a}P_{a}\left( t \right)-\sigma_{a}I_{a}(t),$$

$$\frac{dI_{s}(t)}{dt}=\varepsilon_{s}P_{s}\left( t \right)-\sigma_{s}I_{s}(t),$$

$$\frac{dR\left( t \right)}{dt}=\sigma_{a}I_{a}\left( t \right)+\sigma_{s}I_{s}\left( t \right),$$

where $b$ is the transmission rate; $1/\varepsilon$ is the (mean) pre-infectious period; $1/\sigma$ is the (mean) infectious period; $p$ is the proportion of not presenting symptoms. The basic reproduction number of this model can be derived from the parameters: $R_{0}=p\frac{b}{\sigma_{a}}+\left( 1-p \right)\frac{b}{\sigma_{s}}$.

**Supplementary Note 3: Simulation of screening SARS-CoV-2 cases**

**(1) Daily screening**

***Computation of the proportion of identified cases (not including those infected during the screening period)***

Assuming the school or the office is composed of 1000 individuals, those who did not present symptoms by the time of the first screening (since the epidemic starts), $t_{0}$, $K\left( t_{0} \right)=1000*\left\{ C(t_{0})-B(t_{0}) \right\}$ are recruited to the screening program, where $C(t_{0})$ and $B(t_{0})$ are the cumulative incidence and the proportion of those presented symptoms by $t_{0}$ in the community. Although there are patients infected during the screening period ($1000*\left\{ C\left( t_{0}+T \right)-C(t_{0}) \right\}$) ($T$ is the screening period), they are not counted. In other words, the time of infection $t_{inf}$ is before the screening starts: $t_{inf}<t_{0}$). Those recruited to the screening are categorized into three groups depending on whether they are identified and how they are identified:

1) $D(t)$: the number of cases identified by antigen tests (not by presence of symptoms) at epidemic time $t$ (when tests are performed). Those are identified because the measured viral load is above the detection limit (i.e., viral load of case $i$ at time $t$ $\hat{V}_{i}\left( t \right)>d$, where $d$ is the detection limit). The time $t$ is before the symptom onset (i.e., $t_{inf, i}+g_{i}>t$, where $t_{inf, i}$ and $g_{i}$ are the time of infection and incubation period for case $i$) (Note: For convenience, the time scale of $V\left( t \right)$ in this section is “the time since epidemic start”, not “the time since infection”. Therefore, $V\left( t \right)=\hat{V}\left( t \right)=0$ if they are not infected by the time $t$.)
2) $Y(t)$: the number of cases identified by presence of symptoms (not by tests) at time $t$. The measured viral load is below the detection limit (i.e., $\hat{V}_{i}\left( t=t_{inf, i}+g_{i} \right)<d$).
3) $U(t)$: the number of total unidentified cases by the time $t$.

Thus, the proportion of identified cases at time $t$ by the antigen test or symptom presence, $L\left( t \right)$, is be calculated as follows:

$$L\left( t \right)=\frac{\sum_{{\tau=t}_{0}}^{t} \left( D\left( \tau\right)+Y\left( \tau\right) \right)}{K(t_{0})} .$$

***Computation of the proportion of identified cases (including those infected during the screening period)***

Considering those infected during the screening period, both denominator and numerator of $L\left( t \right)$ changes during the screening period: $K\left( t_{0},t \right)=1000*\left\{ C\left( t \right)-B(t_{0}) \right\}$. $D\left( \tau\right)$ and $Y\left( \tau\right)$ also include those infected cases during the screening period.

**(2) Different screening scenarios with high sensitivity antigen tests**

Screening with antigen tests are performed four times with different schedules over the screening period (10 days): (1) everyday (Day 0, 1, 2, 3), (2) two days interval (Day 0, 2, 4, 6), (3) three days interval (Day 0, 3, 6, 9), (4) late screening (Day 0, 8, 9, 10), and (5) random screening (the timing of test is determined randomly over the 10 days) (**Fig. 2A**). Therefore, the proportion of identified cases at time $t$ by the antigen test or symptom presence, $L\left( t \right)$, not including those infected during the screening period is be calculated as follows:

$$L\left( t \right)=\frac{\sum_{t_{j}<t} D\left( t_{j} \right)+\sum_{{\tau=t}_{0}}^{t} Y\left( \tau\right)}{K(t_{0})},$$

where $t_{j}$ is the timing of $(j+1)$th test. Again, if those infected during the screening period are also included, both denominator and numerator of $L\left( t \right)$ changes during the screening period: $K\left( t_{0},t \right)=1000*\left\{ C\left( t \right)-B(t_{0}) \right\}$. $D\left( t_{j} \right)$ and $Y\left( \tau\right)$ also include those infected cases during the screening period (for the proportions of identified asymptomatic and pre-symptomatic cases, the denominators are asymptomatic and symptomatic cases, respectively).
